# Characteristics and influence of type 2 diabetes in cirrhosis ascites with spontaneous bacterial peritonitis

**DOI:** 10.1101/2020.09.25.20201798

**Authors:** Lin Zhang, Xiaofei Li, Ying Qi, Yang Zhao, Frank Wang, Hongyan Ma, Xuzhen Qin, Danni Fan, Xiangyi Kong, Zhihong Qi, Xiaoyu Zhang

## Abstract

**Objective:** To investigate the characteristics and influence of type 2 diabetes in hepatocirrhosis ascites complicated with spontaneous bacterial peritonitis(SBP) to support clinical management of this condition.

**Methods:** A retrospective study was conducted to collect patients with hepatocirrhosis ascites with hospitalization from January 1, 2016 to June 30, 2019. The patients were classified according to whether they had type 2 diabetes and SBP. Univariate and multivariate binary logistic regression analysis were performed on the data of the two groups.

**Results:** A total of 214 patients were enrolled in the study, including 121 males and 93 females; 21 diabetics and 193 non diabetics; 119 SBP patients and 95 control subjects. There were 18 cases of SBP in hepatocirrhosis ascites complicated with diabetes, accounting for 85.7%, which was significantly higher than that in the non-diabetic group. The times of ascites, age and hospitalization days of cirrhotic ascites complicated with diabetes were 2 (1-3) times, 74 (60-76) years old and 25 (15-36) days, respectively, which were significantly higher than those in the non-diabetic group, P < 0.05. Multivariate analysis showed that diabetes, the times of hepatocirrhosis ascites, hospitalization days and total bilirubin (TBIL) increased the independent risk factors of SBP in hepatocirrhosis ascites, with OR values of 5.126 (1.358-19.345), 1.949 (1.428-2.660), 1.028 (1.010-1.047), 1.006 (1.001-1.010), respectively (P < 0.05).

**Conclusion:** the patients with hepatocirrhosis ascites complicated with diabetes showed older age, longer hospitalization time, more SBP and more ascites; diabetes mellitus, times of ascites, hospitalization days and TBIL increased the risk of SBP in hepatocirrhosis ascites.

## Background

Since the presence or absence of secondary spontaneous bacterial peritonitis (SBP) in cirrhotic ascites is an important factor affecting the progress of liver disease and treatment effect, early diagnosis and implementation of safe and effective treatment can significantly reduce the mortality related to SBP infection. Therefore, it is very important to identify hepatocirrhosis ascites with SBP.^1^ Especially in hepatocirrhosis ascites without infection symptoms (without clinical symptoms of peritonitis), ascites routine, culture of ascites bacteria, molecular detection and other technologies are needed to identify whether there is secondary SBP. However, because of abdominal puncture is invasive examination, the compliance of patients without infection symptoms is not high. There are few outpatient department for exploratory abdominal puncture, it is difficult to ensure the patients without obvious infection symptom can be identified by ascites test for the presence or absence of SBP.^2,3^

At present, the main research is to find out the routine influencing factors of cirrhosis ascites complicated with SBP through convenient and accessible indicators, so as to find out whether there is SBP and intervene as early as possible, and carry out further unconventional examination and anti-infection treatment.^4^ The management of patients with cirrhosis ascites can improve the detection rate of SBP through the evaluation of routine projects. It is in order to effectively ensure the medical safety and medical quality of this group of patients, which has high feasibility and application value.^5^ However, there is no specific study on the occurrence of SBP in cirrhosis ascites. The purpose of this paper is to describe the characteristics of diabetes in cirrhosis ascites and evaluate the risk assessment of SBP in order to provide early clinical intervention and control the occurrence of disease and improve the prognosis.

## Materials and Methods

### Research data

Retrospective study was used to collect the cases of cirrhosis with ascites during hospitalization in Shanghai public health clinical center from January 1, 2016 to June 30, 2019 through hospital information system (HIS) medical record. The diagnosis of cirrhosis requires imaging or pathological basis and ascites should be based on imaging diagnosis.

### Selection criteria

Inclusion criteria: (1) SBP group (infection group) was defined as the routine polymorphonuclear leukocyte (PMN) count more than 500 * 10 ^ 6 / L in ascites of candidate cases; (2) the control group (non infection group) was defined as the routine PMN count was lower than 100 * 10 ^ 6 / L and ascites bacterial culture was negative. Exclusion criteria: Cases of secondary peritonitis, tuberculosis and tumor. Informed consent of the patient was obtained for all diagnostic and treatment activities.

### Research indicators and detection

A retrospective study was conducted to review the case information and data of HIS, LIS and RIS, and to analyze the baseline conditions of the risk group and the control group statistically. The indicators are including hypertension, diabetes, cardiovascular disease, chronic obstructive pulmonary disease, chronic nephrosis, hepatic encephalopathy, gastrointestinal hemorrhage, times of hepatic ascites, age, hospitalization days, peripheral blood white blood cells (WBC), hemoglobin (Hgb), neutrophils (Neu), lymphocyte (LYM), platelet (PLT), alanine aminotransferase (ALT), aspertate aminotransferase (AST), total bilirubin (TBIL), albumin (propagated) and prealbumin (PA), total cholesterol (TC), triglyceride (TG), urea (BUN), creatinine (Cr), prothrombin time (PT), Prothrombin activity (PTA).The above indicators are the first data within 24 hours of admission.

### Statistical analysis

SPSS16.0 software was used for statistical analysis of the research data. Person chi square test and paired chi square test were used for counting data. Through single factor analysis, the indexes with P value < 0.05 were included in multivariate analysis, and logistic regression was used for multivariate analysis. P value < 0.05 had statistical significance.

## Results

### Baseline

In this study, 214 cases of ascites due to cirrhosis were screened, including 121 males and 93 females. The average age was 63 (53-72) years. The average hospitalization days was 15 (10-30) days. the disease was mainly viral hepatitis, with 104 cases of cirrhosis induced by hepatitis B virus or hepatitis C virus, accounting for 50.0%. Child-pugh scores were mainly distributed in Grade B or C, with a total of 196 cases, accounting for 91.6%. SBP occurred in 119 cases, accounting for 55.6%; There were 21 cases of diabetes, accounting for 9.8%. The specific details of the research indicators are shown in **Table1**.

**Table1:** Baseline characteristics of the patients.

| Indicators | NO.of cases(%) |
| --- | --- |
| Cases | 214 |
| Age | 63(53-72) |
| Gender,Male(%) | 121(56.5) |
| SBP(%) | 119(55.6) |
| Child-Pugh A (%) | 18(8.4) |
| Child-Pugh B (%) | 121(56.5) |
| Child-Pugh C (%) | 75(35.0) |
| Comorbidity |  |
| Hypertension(%) | 39(18.2) |
| Diabetes(%) | 21(9.8) |
| CVD(%) | 17(7.9) |
| COPD(%) | 4(1.9) |
| CKD(%) | 11(5.1) |
| HE(%) | 9(4.2) |
| GIB(%) | 38(17.8) |
| Times of hepatocirrhosis ascites | 2(1-3) |
| Hospitalization days | 15(10-30) |
| Etiology |  |
| Viral hepatitis B or C (%) | 107(50.0) |
| Autoimmune (%) | 23(10.7) |
| Schistosomiasis (%) | 27(12.6) |
| Alcoholic (%) | 2(0.9) |
| Hybridism | 11(5.1) |
| Unknown (%) | 44(20.6) |

\* P<0.05 as statistically significant

### Characteristics and influence of diabetes in cirrhosis ascites

In the study group, there were 18 cases of SBP in cirrhotic ascites complicated with diabetes, accounting for 85.7%, which was significantly higher than that in the non-diabetic group. The times of ascites, age and hospitalization days of cirrhotic ascites complicated with diabetes were 2 (1-3) times, 74 (60-76) years old and 25 (15-36) days respectively, which were significantly higher than those in the non-diabetic group; all P < 0.05. Gender, Child-Pugh classification, hypertension, cardiovascular disease, chronic obstructive pulmonary disease, chronic nephrosis, hepatic encephalopathy, gastrointestinal hemorrhage, peripheral blood white blood cell (WBC), Hemoglobin (Hgb), Neutrophil (neu), Lymphocyte (lym), Platelet (PLT), Alanine aminotransferase (ALT), Aspartate aminotransferase (AST), Total Bilirubin (TBIL), albumin (ALB), prealbumin (PA), total cholesterol (TC), triglyceride (TG), urea (BUN), creatinine (Cr), Prothrombin Time (PT), Prothrombin Activity (PTA) had no significant difference between the two groups, P > 0.05. The specific details of the research indicators were shown in **Table2**.

**Table-2:** Characteristics and influence of diabetes in hepatocirrhosis ascites.

| **Indicators** | **Diabetes** | **Non-Diabetes** | **P Value** |
| --- | --- | --- | --- |
| Cases | 21 | 193 |  |
| Age | 74(60-76) | 62(52-71) | 0.018* |
| Gender, Male(%) | 10(47.6) | 111(57.5) | 0.385 |
| SBP(%) | 18(85.7) | 101(52.3) | 0.003* |
| Child-Pugh A (%) | 1(47.6) | 17(8.8) | 1.000 |
| Child-Pugh B (%) | 16(4.8) | 105(54.4) | 0.056 |
| Child-Pugh C (%) | 4(19.0) | 71(36.8) | 0.106 |
| Comorbidity |  |  |  |
| Hypertension(%) | 5(23.8) | 34(17.6) | 0.550 |
| CVD(%) | 3(14.3) | 14(7.3) | 0.224 |
| COPD(%) | 0(0) | 4(2.1) | 1.000 |
| CKD(%) | 1(4.8) | 10(5.2) | 1.000 |
| HE(%) | 0(0) | 9(4.7) | 0.604 |
| GIB(%) | 5(23.8) | 33(17.1) | 0.546 |
| Times of hepatocirrhosis ascites | 2(1-3) | 1(1-2) | 0.039* |
| Hospitalization days | 25(15-36) | 14(9-29) | 0.011* |
| Blood test |  |  |  |
| WBC($10^9/L$) | 3.8(2.8-5.1) | 4.4(3.1-6.3) | 0.201 |
| Hgb(g/L) | 103(62-114) | 106(89-121) | 0.149 |
| Neu($10^9/L$) | 2.67(1.76-3.73) | 2.96(1.85-4.36) | 0.369 |
| LYM($10^9/L$) | 0.58(0.23-1.06) | 0.70(0.49-1.24) | 0.071 |
| PLT($10^9/L$) | 78(52-103) | 67(48-115) | 0.529 |
| ALT(U/L) | 26(17-41) | 28(18-47) | 0.390 |
| AST(U/L) | 36(25-45) | 44(30-69) | 0.075 |
| TBIL($\mu\text{mol/L}$) | 29(14-53) | 36(21-71) | 0.205 |
| ALB(g/L) | 29(27-32) | 30(25-34) | 0.643 |
| PA(mg/L) | 57(29-99) | 51(25-91) | 0.745 |
| BUN($\text{mmol/L}$) | 5.5(4.48-14.50) | 6.09(4.65-9.82) | 0.996 |
| Cr($\mu\text{mol/L}$) | 81.6(67.4-140.1) | 71.2(58.0-102.9) | 0.068 |
| PT(sec) | 16(15.4-17.6) | 16.7(15.3-19.4) | 0.284 |
| PTA(%) | 69(58-74) | 62(49-71) | 0.122 |

\* P<0.05 as statistically significant

### SBP group and control group in cirrhotic ascites

In the infection group, among the SBP cases of cirrhosis ascites, 18 cases were complicated with diabetes, accounting for 15.1%, ascites occurred 2(1-3) times. All of them were significantly higher than the control group. In SBP cases with cirrhosis ascites, ALT, Cr, PT were 30(19-50)U /L, 75.0(62.5-110.9) Umol / L, 17.2(15.8-19.7) SEC, which were significantly higher than those in the control group, P < 0.05. Gender and grade of the Child - Pugh, hypertension, cardiovascular disease, chronic obstructive pulmonary disease, chronic nephrosis, hepatic encephalopathy, gastrointestinal bleeding, peripheral blood white blood cells (WBC), Hemoglobin (Hgb), Neutrophils (Neu), Lymphocyte (LYM), Platelet (PLT), Aspertate aminotransferase (AST), Total bilirubin (TBIL), Albumin (propagated) and prealbumin (PA), Total cholesterol (TC), Triglyceride (TG), Urea (BUN), Prothrombin activity (PTA) in two groups had no significant difference, P> 0.05. The specific details of the research indicators are shown in **Table3**.

**Table-3:** SBP group and control group in cirrhotic ascites.

| **Indicators** | **Infection(SBP)** | **Control(No-SBP)** | **P Value** |
| --- | --- | --- | --- |
| Cases | 119 | 95 |  |
| Age | 63(52-73) | 64(54-71) | 0.453 |
| Gender,Male(%) | 69(58.0) | 52(54.7) | 0.634 |
| SBP(%) |  |  |  |
| Child-Pugh A (%) | 8(6.7) | 10(10.5) | 0.319 |
| Child-Pugh B (%) | 71(59.7) | 50(52.6) | 0.302 |
| Child-Pugh C (%) | 40(33.6) | 35(36.8) | 0.623 |
| Comorbidity |  |  |  |
| Hypertension(%) | 18(15.1) | 21(22.1) | 0.189 |
| CVD(%) | 18(15.1) | 3(3.2) | 0.003* |
| COPD(%) | 8(6.7) | 9(9.5) | 0.460 |
| CKD(%) | 1(0.8) | 3(3.2) | 0.325 |
| HE(%) | 4(3.4) | 7(7.4) | 0.222 |
| GIB(%) | 3(2.5) | 6(6.3) | 0.190 |
| Times of hepatocirrhosis ascites | 20(16.8) | 18(18.9) | 0.684 |
| Hospitalization days | 2(1-3) | 1(1-2) | <0.001* |
| Blood test |  |  |  |
| WBC($10^9/L$) | 21(13-37) | 12(7-20) | <0.001* |
| Hgb(g/L) |  |  |  |
| Neu($10^9/L$) | 4.5(3.1-6.6) | 4.3(3.0-6.1) | 0.371 |
| LYM($10^9/L$) | 106(91-119) | 106(81-124) | 0.548 |
| PLT($10^9/L$) | 2.86(2.00-4.78) | 2.76(1.67-3.87) | 0.171 |
| ALT(U/L) | 0.67(0.46-1.16) | 0.79(0.52-1.30) | 0.091 |
| AST(U/L) | 67(47-97) | 72(48-130) | 0.487 |
| TBIL(umol/L) | 30(19-50) | 25(15-40) | 0.032* |
| ALB(g/L) | 44(29-67) | 40(31-63) | 0.570 |
| PA(mg/L) | 38(21-93) | 31(19-50) | 0.089 |
| BUN(mmol/L) | 30(26-33) | 30(25-34) | 0.744 |
| Cr(umol/L) | 48(26-88) | 61(26-92) | 0.407 |
| PT(sec) | 6.56(4.67-11.25) | 5.69(4.46-9.14) | 0.114 |
| PTA(%) | 75.0(62.5-110.9) | 67.5(50.7-98.1) | 0.023* |
| Child-Pugh A (%) | 17.2(15.8-19.7) | 16.3(14.7-18.3) | 0.002* |
| Child-Pugh B (%) | 60(48-70) | 64(51-73) | 0.211 |

\* P<0.05 as statistically significant

### Correlation analysis of SBP affecting hepatocirrhosis ascites

Univariate analysis showed that diabetes, times of ascites, hospitalization days and total bilirubin (TBIL) were risk factors for SBP in cirrhosis ascites, with OR values of 5.465 (1.559-19.161), 1.683 (1.273-2.225), 1.031 (1.013-1.049), 1.006 (1.001-1.010). Platelet (PLT) was the protective factor of SBP in hepatocirrhosis ascites, OR value was 0.996 (0.99). Gender and grade of the Child-Pugh, hypertension, cardiovascular disease, chronic obstructive pulmonary disease, chronic nephrosis, hepatic encephalopathy, gastrointestinal bleeding, peripheral blood white blood cells (WBC), Hemoglobin (Hgb), Neutrophils (Neu), Lymphocyte (LYM), Aspertate aminotransferase (AST), Total bilirubin (TBIL), Albumin (ALB) and prealbumin (PA), Total cholesterol (TC), Triglyceride (TG), Urea (BUN), Prothrombin activity (PTA) did not affect the risk of SBP in cirrhosis ascites (P > 0.05).

Multivariate analysis showed that diabetes, times of cirrhosis ascites, hospitalization days and TBIL were the independent risk factors for SBP in cirrhosis ascites, with OR values of 5.126 (1.358-19.345), 1.949 (1.428-2.660), 1.028 (1.010-1.047), 1.006 (1.001-1.010), P < 0.05. Platelet (PLT) was a confounding factor which did not affect the incidence of cirrhosis ascites with SBP, P > 0.05. More details of research indicators were in **Table4**.

**Table-4:**
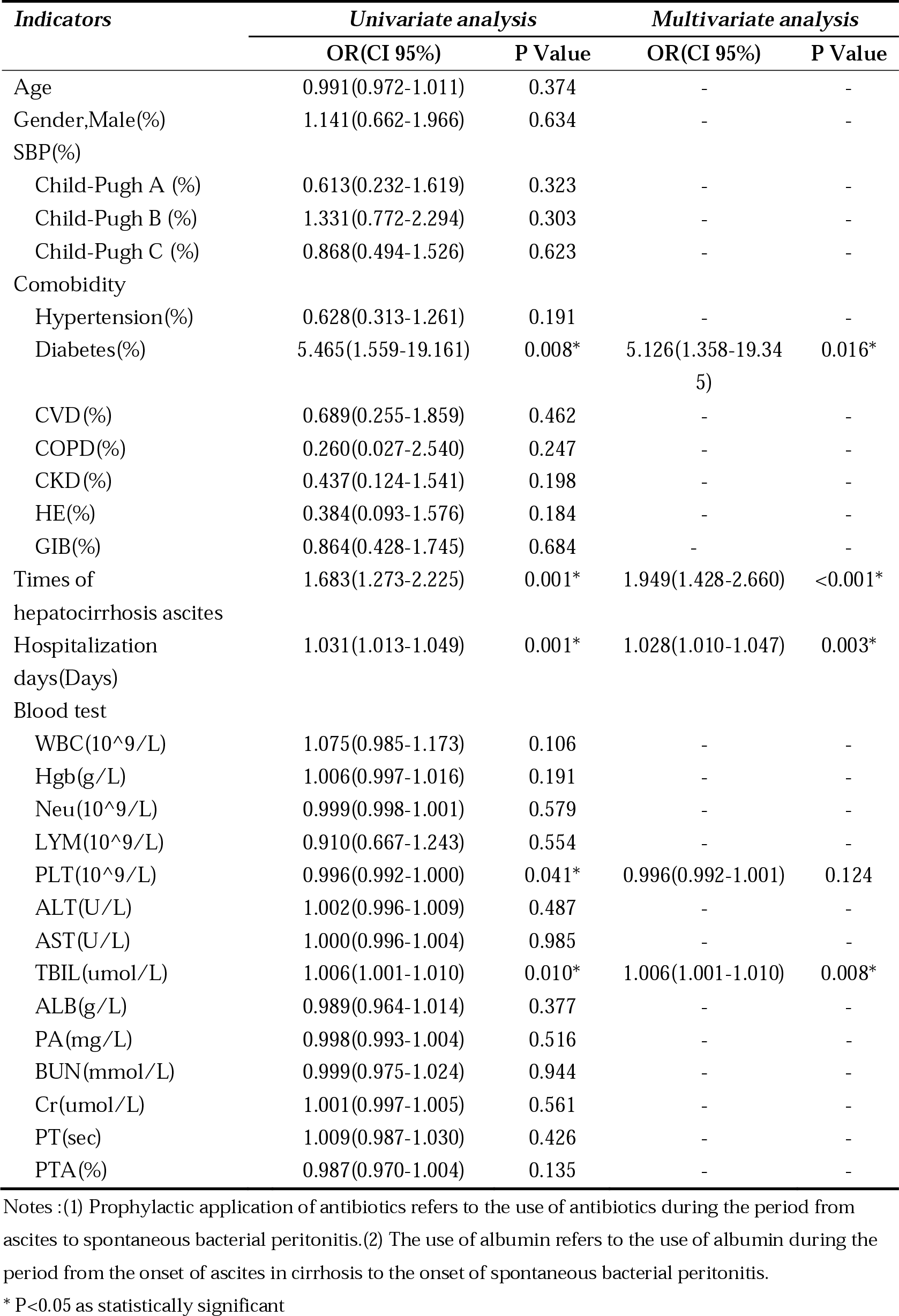
Correlation factor strength analysis of SBP.

| **Indicators** | **Univariate analysis** |  | **Multivariate analysis** |  |
| --- | --- | --- | --- | --- |
|  | **OR(CI 95%)** | **P Value** | **OR(CI 95%)** | **P Value** |
| Age | 0.991(0.972-1.011) | 0.374 | - | - |
| Gender,Male(%) | 1.141(0.662-1.966) | 0.634 | - | - |
| SBP(%) |  |  |  |  |
| Child-Pugh A (%) | 0.613(0.232-1.619) | 0.323 | - | - |
| Child-Pugh B (%) | 1.331(0.772-2.294) | 0.303 | - | - |
| Child-Pugh C (%) | 0.868(0.494-1.526) | 0.623 | - | - |
| Comorbidity |  |  |  |  |
| Hypertension(%) | 0.628(0.313-1.261) | 0.191 | - | - |
| Diabetes(%) | 5.465(1.559-19.161) | 0.008* | 5.126(1.358-19.345) | 0.016* |
| CVD(%) | 0.689(0.255-1.859) | 0.462 | - | - |
| COPD(%) | 0.260(0.027-2.540) | 0.247 | - | - |
| CKD(%) | 0.437(0.124-1.541) | 0.198 | - | - |
| HE(%) | 0.384(0.093-1.576) | 0.184 | - | - |
| GIB(%) | 0.864(0.428-1.745) | 0.684 | - | - |
| Times of hepatocirrhosis ascites | 1.683(1.273-2.225) | 0.001* | 1.949(1.428-2.660) | <0.001* |
| Hospitalization days(Days) | 1.031(1.013-1.049) | 0.001* | 1.028(1.010-1.047) | 0.003* |
| Blood test |  |  |  |  |
| WBC($10^9/L$) | 1.075(0.985-1.173) | 0.106 | - | - |
| Hgb(g/L) | 1.006(0.997-1.016) | 0.191 | - | - |
| Neu($10^9/L$) | 0.999(0.998-1.001) | 0.579 | - | - |
| LYM($10^9/L$) | 0.910(0.667-1.243) | 0.554 | - | - |
| PLT($10^9/L$) | 0.996(0.992-1.000) | 0.041* | 0.996(0.992-1.001) | 0.124 |
| ALT(U/L) | 1.002(0.996-1.009) | 0.487 | - | - |
| AST(U/L) | 1.000(0.996-1.004) | 0.985 | - | - |
| TBIL(umol/L) | 1.006(1.001-1.010) | 0.010* | 1.006(1.001-1.010) | 0.008* |
| ALB(g/L) | 0.989(0.964-1.014) | 0.377 | - | - |
| PA(mg/L) | 0.998(0.993-1.004) | 0.516 | - | - |
| BUN(mmol/L) | 0.999(0.975-1.024) | 0.944 | - | - |
| Cr(umol/L) | 1.001(0.997-1.005) | 0.561 | - | - |
| PT(sec) | 1.009(0.987-1.030) | 0.426 | - | - |
| PTA(%) | 0.987(0.970-1.004) | 0.135 | - | - |

Notes : (1) Prophylactic application of antibiotics refers to the use of antibiotics during the period from ascites to spontaneous bacterial peritonitis. (2) The use of albumin refers to the use of albumin during the period from the onset of ascites in cirrhosis to the onset of spontaneous bacterial peritonitis.
\* P<0.05 as statistically significant

## Discussion

This study was focused on the characteristics and influence of diabetes mellitus in cirrhotic ascites complicated with spontaneous bacterial peritonitis. In order to avoid the influence of other factors, univariate and multivariate analysis methods were used to exclude confounding factors and determine the independent correlation. Due to previous reports, some factors that may affect the occurrence of spontaneous bacterial peritonitis were also included in the statistical analysis, such as the prophylactic use of antibiotics and human albumin supplementation in some populations.

The study found that the risk of spontaneous bacterial peritonitis increased with the frequency of cirrhosis ascites. Therefore, early intervention to avoid or reduce complications can effectively reduce the occurrence of spontaneous bacterial peritonitis.

In this study, the view that serum bilirubin was the independent influencing factor of cirrhosis ascites with SBP is in accordance with the present study reports the mainstream. Studies have shown that serum bilirubin is independently related to SBP in cirrhotic ascites patients, and is an independent influencing factor for spontaneous bacterial peritonitis.^6-8^ However, patients with liver cirrhosis often show hyperbilirubinemia due to bile duct compression and deformation or related inflammatory infiltration. It is difficult to infer that it is complicated by SBP. In any case, from the perspective of prevention and treatment, whether it is liver itself or bacterial inflammation, early clinical intervention of hyperbilirubinemia is of great significance.

In this study, diabetes mellitus was an independent influencing factor for cirrhosis ascites with SBP, which was consistent with most current research reports.^9-11^ Thereby, for patients with cirrhosis, how to control blood glucose clinically and to what extent to minimize its impact on cirrhosis ascites with SBP is the key project of the next research.

What is inconsistent with other studies in this study is that Child-pugh score and albumin are not related to the independent correlation of cirrhosis ascites with SBP. However, Zuwala-Jagiello J et al. believed that Child-pugh score and albumen were independent risks of SBP in cirrhotic ascites.^12^ The main reason is that some researchers have done this research report in the early stage, and they have obtained this information clinically and have carried out early intervention. In our study, there were 168 cases of prophylactic intervention, including 78.2% in SBP group and 78.9% in control group. Albumin was used in 127 cases, 60.0% in SBP group and 58.9% in control group. From now on, we or other research teams can further verify the effect of prophylactic application of antibiotics or human serum albumin on reducing the incidence of spontaneous bacterial peritonitis in cirrhotic ascites, and whether Child-Pugh score and albumin are the related influencing factors of SBP in cirrhotic ascites.

This study found that platelets were not independently associated with cirrhosis ascites with spontaneous bacterial peritonitis, but were confounding factors. Metwally K reported that thrombocytopenia was an independent risk of cirrhotic ascites with SBP.^13^ This is not consistent with our study, which may be related to our early prophylactic use of antibiotics and human albumin in at-risk cases. However, Metwally K reports that there is no explanation for the prophylactic use, so that it is hard to identify the reasons for the differences.

In our study, we described the clinical characteristics of diabetes mellitus in cirrhotic ascites with SBP. At the same time, the occurrence of SBP caused by diabetes mellitus was further confirmed to be highly correlated with cirrhotic ascites, and the frequency of ascites occurrence, hospitalization days and bilirubin level were still closely correlated with SBP.

After a rough assessment, it was found that under some preventive interventions, the influential factors reported in some previous studies were not obvious in cirrhotic ascites with SBP, which indicated that the importance of further evaluation for the influential factors.

Meanwhile, there are a number of limitations in this study. The type, dose and course of prophyllactic antibiotics and the dosage and method of human albumin were not included in the statistical analysis. Moreover, our study did not define the onset time of diabetes mellitus, and its effect on SBP was only the difference in the intensity of association.

In conclusion, with the in-depth study of cirrhotic ascites, the scope and degree of clinical intervention on risk factors are increased, previous evaluation items and evaluation models may change at different levels, and further analysis, evaluation and application are needed in clinical studies. The present study suggests that diabetes mellitus, recurrent ascites, prolonged hospitalization, and high bilirubin levels are still risk factors for SBP in cirrhotic ascites. The key points of clinical intervention for cirrhotic ascites with SBP are to control diabetes, avoid ascites recurrence, reasonably arrange hospitalization time and reduce bilirubin level.

## Data Availability

The study was supported by the Shanghai Public Health Clinical Center for data access.

## Contributions

Conceptualization, Xiao-Yu Zhang, Lin Zhang; Formal analysis, Xiao-Yu Zhang; Methodology, Xiao-Yu Zhang, Lin Zhang; Writing – original draft, Xiao-Yu Zhang, Lin Zhang; Writing – review & editing, all authors. All authors have read and agreed to the published version of the manuscript.

## Acknowledgments

The study was supported by the Shanghai Public Health Clinical Center for data access.

## Ethical approval

All the data received Institutional Review Board (IRB) approval by the Ethics Committee of the Shanghai Public Health Clinical Center.

